# CD16a^high^ NK cell infiltration and spatial relationships with T cells and macrophages can predict improved progression-free survival in high grade ovarian cancer

**DOI:** 10.1101/2021.06.08.21258566

**Authors:** Sarah Nersesian, Stacey N. Lee, Stephanie Grantham, Liliane Meunier, Laudine Communal, Thomas Arnason, Dirk Arnold, Brad H. Nelson, Anne-Marie Mes-Messon, Jeanette E. Boudreau

## Abstract

**Background:** High grade serous cancer (HGSC) remains a highly fatal malignancy with less than 50% of patients surviving 5 years after diagnosis. Despite its high mutational burden, HGSC is relatively refractory to checkpoint immunotherapy, suggesting that additional features of the cancer and its interactions with the immune system remain to be understood. Natural killer (NK) cells may contribute to HGSC control, but the role(s) of this population or its subsets in this disease are poorly understood.

**Methods:** We used a TMA containing duplicate treatment-naïve tumors from 1145 patients with HGSC and a custom staining panel to simultaneously measure macrophages, T cells and NK cells, separating NK cells based on CD16a expression. Using pathologist-validated digital pathology, machine learning, computational analysis and Pearson’s correlations, we quantitated infiltrating immune cell density, co-infiltration and co-localization with spatial resolution to tumor region. We compared the prognostic value of innate, general, and adaptive immune cell “neighborhoods” to define characteristics of HGSC tumors predictive for progression-free survival and used flow cytometry to define additional features of the CD16a^dim^ NK cell subset.

**Results:** NK cells were observed in >95% of tumor cores. Intrastromal localization of CD16a^low^ and CD16a^high^ NK cells was associated with shorter and longer progression-free survival, respectively. CD16a^high^ NK cells most frequently co-localized with T cells and macrophages; their proximity was termed an “adaptive” neighborhood. We find that tumors with more area represented by adaptive immune cell neighborhoods corresponded to superior progression free survival. In contrast, CD16a^low^ NK cells did not co-infiltrate with other immune cell types, and expressed the ectonucleotidases, CD39 and CD73, which have been previously associated with poor prognosis in patients with HGSC.

**Conclusions:** Progression-free survival for patients with HGSC may be predicted by the subset of NK cells within the tumor infiltrate (i.e. CD16a^high^ vs. CD16a^low^). NK cell subtypes were associated predictable co-infiltrating and co-localizing leukocyte subsets, suggesting that their presence and activity may influence, or be influenced by the tumor microenvironment. Our data suggest that immunotherapeutic strategies for HGSC should consider the constitution of NK cell subsets and may benefit from mobilizing and activating CD16^high^ NK cells.

## BACKGROUND

High grade serous carcinomas (HGSC) are the most common, aggressive and genetically unstable form of ovarian cancer, and associated with the poorest 5-year survival rate[1, 2]. A lack of effective screening methods and specific signs and symptoms typically leads to late-stage diagnosis, when patients frequency have metastatic disease (80%, stage III or IV)[1, 2]. Although tumors initially respond to a combination of platinum and taxane based chemotherapy, over 70% recur, most as chemotherapy resistant[2, 3]. Despite a high mutational burden and consequently high neoantigen load, anti-PD-L1 therapies have been largely ineffective against HGSC (overall response rate 0-11.5%)[4-6]. Perhaps the dynamic nature of the tumor, and loss of human leukocyte antigen (HLA) expression challenges the generation and efficacy of anti-tumor (HLA-restricted) T cell responses[7, 8]. Collaboration from other immune cell types, including natural killer (NK) cells could extend immune reactivity against HGSC; these cells has not been as extensively studied.

NK cells comprise approximately 10% of circulating cells and were first defined based on their antitumor potential[9]. NK cells differentiate from the common lymphoid progenitor in the bone marrow and are most closely related to T cells with similar features for cytokine production and cytotoxicity of target cells. The potent anti-tumor capability of NK cells is antigen-independent[10-12]. NK cells can be broadly categorized into two main populations based on CD16a and CD56 expression[10]. CD56^bright^/CD16a^low^ NK cells primarily produce cytokines, such as IFN-γ, TNF and GM-CSF[13]. CD56^dim^/CD16a^high^ NK cells can also produce these cytokines, and additionally are the primary cytotoxic effectors, through expulsion of perforin and granzymes, or via the Fas and TRAIL death-receptor pathways[10, 14]. NK cells exhibit an array of phenotypes and related functions, which are governed by variable expression and co-expression of germline-encoded receptors[15]. The functions of NK cells are diverse, ranging from missing self-reactivity (an ability to kill cells based on loss of self HLA), induced-self reactivity (killing cells with over-expression of stress-associated ligands), antibody-dependent cellular cytotoxicity and immunoregulation[16, 17].

Despite significant suppression within the tumor microenvironment (TME) and relatively low numbers of NK cells infiltrating tumors, two studies conducted with a small number of HGSC core biopsies (<300 patients) found infiltrating NK (CD57^+^) cells to be associated with improved clinical outcomes[18, 19]. Interestingly, these associations were strengthened when considering NK cells in the intraepithelial (IE) region of the tumor[18, 19]. This is particularly relevant given that NK cells existing within the cancer cell nest (IE region) express increased levels of inhibitory receptors, including NKG2A, compared to those in the stromal region, suggesting a higher propensity for inhibition compared to activation[20]. These studies provide the important proof-of-concept that NK cell infiltration is associated with beneficial outcomes clinically, but CD57 is only expressed on a relatively mature subset of NK cells and therefore may not accurately represent the entire NK cell population[18, 21-23]. To enable analysis of the *entire* infiltrating NK cell population, we designed and validated a multiplex panel for precise NK cell identification in HGSC by combining antibodies against CD16a (NK cells, macrophages), CD94 (a component of the binding complexes formed by the NK receptor group 2 (NKG2) receptors present in some form on all NK cells, T cells), CD3 (T cells) and CD68 (macrophages and mononuclear phagocytes) and pancytokeratin[24, 25]. With this panel, we find that NK cells infiltrated the majority of HGSC cores, but we observed subtype specific patterns: CD16a^low^ NK cells typically infiltrated alone, while CD16a^high^ NK cells more often co-infiltrated with T cells and macrophages. This latter “adaptive” group was associated with improved progression-free survival among patients with HGSC, while a relative enrichment for “innate” CD16a^low^ areas was associated with shorter progression free survival time. CD16a^high^ NK cell infiltration was further associated with stage 4 and grade 3 tumors, and adaptive neighborhoods were more often found in tumors harboring *BRCA1/2* mutations. CD16a^low^ NK cells correspond with CD39/73 expression, and were associated with features of immunoregulation and poorer prognosis. Taken together, our findings underscore a positive prognostic impact of CD16^high^ NK cells, and support strategies to maximize the antitumor activities of this population in patients with HGSC.

## METHODS

### Patients and Tumor Microarrays

We conducted multiplex immunofluorescence staining followed by quantitative analysis on a formalin-fixed paraffin-embedded (FFPE) HGSC tumor microarray (TMA). The TMA slides were obtained after scientific review from the COEUR (The Canadian Ovarian Experimental Unified Resource) committee. Patient samples were collected from 12 Canadian ovarian cancer biobanks, from 1992 to 2014, and detailed patient demographics and tumor characteristics have been published previously[26]. The COEUR TMA consists of duplicate core tumors isolated from 1159 patients with HGSC. Following tissue staining, image processing, and pathologist review (detailed below), 1145 patients were analyzed. All methods for specimens and clinical information collection and subsequent analyses were approved by Dalhousie University’s Research Ethics Board (#2020-5060). The use of primary peripheral blood mononuclear cells (PBMC) to validate antibodies were approved by the Dalhousie University REB (#2016-3842) and the Canadian Blood Services REB (#2016-016) and collected in collaboration with the Canadian Blood Services Blood4Research program.

### OPAL Multiplex Immunofluorescence

Our multiplex immunofluorescence protocol was established in accordance with the Society for Immunotherapy of Cancer’s best practices for multiplex immunohistochemistry and immunofluorescence staining and validation[27]. TMA slides were de-paraffinized, rehydrated, and then fixed in 10% neutral buffered formalin for 25 minutes. Antigen retrieval was conducted by microwave treatment (2 minutes at 100% power followed by 15 minutes at 20% power, 1000W microwave) in Tris-EDTA buffer (pH 9). Slides were then cooled for 15 minutes at room temperature, rinsed with deionized water, then with Tris-Buffered Saline and Tween 20 (TBST) buffer. The tyramide signal amplification (TSA)-based IF staining protocol was conducted according to the Opal 7-colour manual IHC kit (Akoya Bio, cat#NEL811001KT). Slides were incubated in blocking buffer for 10 minutes (Akoya Bio) to stabilize epitopes and reduce background staining). Slides were then incubated with selected primary antibody (summarized below) for 45 minutes, rinsed in TBST, and incubated with secondary anti-mouse and anti-rabbit HRP (Akoya Bio) for 10 minutes. Fluorophore staining was then conducted with a selected Opal fluorophore in Opal amplification diluent (Akoya Bio, cat#FP1498). The slides were then rinsed and underwent second microwave treatment to remove the previous antibody. Steps were sequentially repeated for each antibody in the multiplex panel (Supplementary Table 2). The order of antibody staining was optimized empirically, and generally occurred in the order from most to least frequent epitope in the panel (Supplementary Table 2). Following the staining of the last antibody, slides were incubated with Spectral DAPI (Akoya Bio) for 5 minutes, rinsed and mounted in Prolong Gold mounting media (Invitrogen). Cell surface specific antibody binding patterns were validated by a pathologist (TA) (Supplementary Figure 2).

### Multispectral Analysis and Automated Quantitative Pathology

Images were captured using a Mantra 2 Quantitative Pathology Workstation (Akoya Bio), at 20x magnification, with multispectral separation capabilities at 10 nm wavelength intervals from 420 nm to 740 nm. Images were then visualized and processed in InForm Tissue Finder Software (Akoya Bio, cat#CLS135783) to conduct multispectral analysis (extracting fluorescent signatures) and subsequent quantitative pathology. Monoplexed stains were used to calibrate spectral imaging, creating a spectral reference library, as described by manufacturer’s instructions (Akoya Bio). Following spectral unmixing, tissues were segmented based on cytokeratin staining and autofluorescence to define regions as tumor epithelia, tumor stroma, vasculature/autofluorescence or off the tumor core. This was followed by cellular segmentation which identified individual nuclei based on DAPI staining. Each cell was then assigned a unique cell ID which was used to quantify the antibody signal surrounding the individual nuclei. These antibody signatures were used to phenotype cells into epithelial cells (PanCK^+^CD^-^), stromal cells (PanCK^-^CD^-^), CD16a^high^ NK cells (CD16a^high^CD94^+/-^CD68^-^CD3^-^), CD16a^low^ NK cells (CD16a^low^CD94^+^CD68^-^CD3^-^), T cells (CD3^+^), Macrophages (CD68^+^) or strongly auto-fluorescent red blood cells (PanCK^-^CD^-^ Autofluorescent^++^). All digital scoring and quantification were validated by TA (Supplementary Figure 3). Consistency between cores were also assessed and validated (Supplementary Figure 4).

### Data Processing

Data were organized and processed through phenoptR Reports in R Studio. Cell segmentation phenotyping data from 2318 cores were exported as a summary of each phenotype density (cells/mm^2^). Of the 2318 cores, 15 (0.65%) were lost during tissue processing and/or staining, 56 (2.4%) and 9 (0.39%) cores were excluded because they did not reach our pre-determined requisite threshold of 1000 cells/mm^2^ in their epithelial and stromal compartments respectively. Excluded cores represented a total of six patients. When our algorithm reported cell counts >1.5 interquartile ranges above the third, or below the first quartile, they were manually reviewed. These outliers included 52 (2.24%) CD3+, 8 (0.34%) CD68+, 67 (2.9%) CD16a^high^ and 76 (3.3%) CD16a^low^cores. Of these, 83 (3.6%) were identified to have artifacts, intrusive autofluorescence or unspecific staining leading to incorrect phenotyping, and therefore were removed. Following this, 2058 cores representing 1145 patients were included in our analysis.

### Spatial Pathology Analysis

Using the cellular x,y coordinates and associated phenotype obtained from InForm quantitative pathology, each cell was used as starting points for radial spatial pathology. In pilot studies, we determined that cell-cell impacts could be best approximated by 22µm distance (approximately 2 lymphocyte widths). The following code was developed in MatLab to identify, within 22µm distance, the fraction of cells of the core, that fit within each pre-defined neighborhood: Adaptive (at least one of each T cell, Mac and CD16a^high^ NK cell, Neighborhood A), Innate (at least two CD16a^low^ NK cell, Neighborhood B) or a General (any three immune cells, Neighborhood C). These neighborhoods, dictated by our co-infiltration studies, are described in additional detail below. The following coding strategy was used to identify how many cells fell within each of the three clusters.

We used a parameter $R>0$ to determine spatial proximity and define the following three types of regions:

> \begin(itemize)
>
> \item Neighborhood A consists of those points of the image plane where at least one
>
> \textt(CD68+ Macrophages) cell, one \texttt(CD16a^high^ NK cells) and one \texttt(CD3+ T cells) are within a radius of $R$.
>
> \item Neighborhood B consists of those points of the image plane where at least two
>
> \texttt(CD16a^low^ NK cells) cells are within a radius of $R$.
>
> \item Neighborhood C consists of those points of the image plane where at least three immune cells are within a radius of $R$, irrespective of their phenotype.
>
> \end(itemize)

In order to determine the fraction of immune cell neighborhoods in the \texttt(Stromal CD-) and \texttt(Epithelial CD-) regions, we create one two-dimensional map for each type of immune cell by adding binary valued radial basis functions of width $R$ centred at each immune cell. Superimposing those maps according to the neighborhoods outlined above (A, B, C), enabled calculation of binary values reflecting whether or not the defined immune cell type exists within the same region. Two-dimensional interpolation allows querying those maps at the locations of all \texttt(Stromal CD-) and \texttt(Epithelial CD-) cells.

### Cell Lines

Human cell lines used in flow cytometry assays included the HLA class I^-^ K562 (American Type Culture Collection (ATCC)), and the ovarian cancer cell lines OVCAR4 (kindly provided by the laboratory of Dr. Barbara Vanderhyden) and OVCAR5. All cells were cultured at 37°C in 5% CO_2_ in RPMI-1604 media with 10% heat inactivated FBS, 100 U/ml penicillin, and 100 mg/ml streptomycin sulfate.

### Flow Cytometry and Gating Strategy

Isolated human PBMCs were stimulated using K562, OVCAR-4, and OVCAR-5 cells (3:1 ratio of PBMC:Target cell) for 5 hr in the presence of anti-CD107a (H4A3, BD Biosciences). Non-specific antibody binding was blocked by addition of 0.5 mg/mL human Fc block (BD Biosciences). Viable human NK cells were identified by excluding dead cells and gating for human CD56^+^CD3^-^ cells (live/dead fixable dye, BD Biosciences). All antibodies, clones, and sources used to identify NK cell subsets are shown in supplementary table 3. Fluorescence was measured using a BD FACS Symphony (BD Biosciences) and analysed using FlowJo 10 software (FlowJo LLC), gating strategy is noted in supplementary figure 4.

### Statistical Analysis

GraphPad Prism 8 was used to generate all graphically visualized data. Specifically, Kaplan-Maier survival curves followed by log-rank testing were used to determine significant differences between various groups. High and low infiltrated tumors were defined based on median infiltration density of each immune cell (cells/mm^2^). Differences in infiltration numbers between groups categorized by clinical parameters were determined by one or two-way ANOVAs, as appropriate. To evaluate the co-infiltration between intratumoral immune cells or neighborhoods, a Pearson’s correlation was conducted with all r values listed for each pair in the correlation matrix. The threshold for statistical significance was set at alpha = 0.05; however, non-statistical but clinically relevant associations, defined by the authors, were described along with corresponding *p* values.

## RESULTS

### NK cells are frequently found within HGSC tumors and demonstrate superior ability to infiltrate IE regions compared to T cells and macrophages

We developed a novel Opal multiplex IF panel to accurately identify and capture immune cell populations within HGSC tissues with a focus on NK cells. To circumvent the challenges associated with epithelial cell expression of CD56 that have been reported in ovarian cancer[21], we used a combination of CD94 and CD16a to capture both the CD16^high^ and CD16^low^ populations of NK cells, which were further defined as CD3^-^CD68^-^ (Figure 1A). T cells and macrophages were defined as CD3^+^CD16^-^CD94^+/-^ and CD68^+^CD16^+/-^CD94^-^, respectively. Staining and subsequent digital pathology, including tissue and cellular segmentation, was validated by TA. Automated tissue segmentation was highly concordant with manual pathology evaluation, while cellular segmentation was comparable. We found that InForm software slightly underreported (∼84% of actual) cell counts in IE and overcounted (∼108% of actual) in IS regions (Supplementary Figure 2). We used this panel along with spectral unmixing and digital pathology to quantify the number of each immune cell population by HGSC core (Figure 1B).

**Figure 1:**
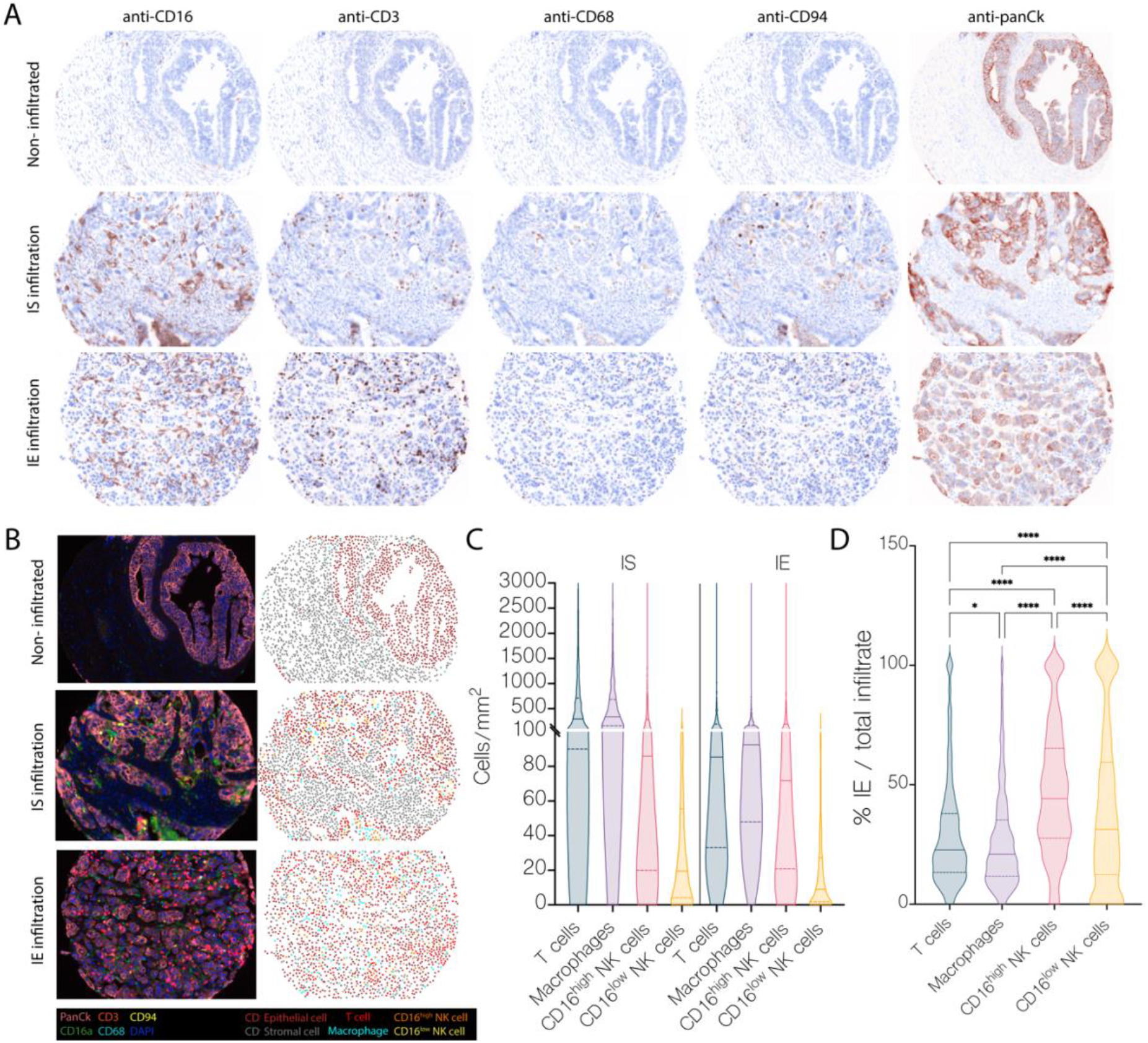
Identification and quantification of NK cells, T cells and macrophages in HGSC TMA by custom OPAL multiplex IF panel reveals preferential recruitment of CD16a^high^ NK cells to the epithelial region. A) Representative pathology views of each individual marker in our TMA: CD16a, CD3, CD68, CD94 and pancytokeratin (panCk) for a non-immune infiltrated, IS infiltrated and IE infiltrated tumor cores; and B) corresponding multiplex overlay (left: anti-CD16a (green), anti-CD3 (red), anti-CD68 (light blue), anti-CD94 (yellow), anti-panCk (pink) and DAPI (dark blue)), with phenotyping map (right: stromal cell (gray), epithelial cell (maroon), T cell (red), Macrophage (blue), CD16a^high^ NK cell (orange), CD16a^low^ NK cell (yellow)). C) Violin plots, with median noted by the solid line, demonstrate the total range of cell density, as a measure of cell count over area (cells/mm^2^) by tumor compartment (IS vs. IE). D) Violin plots representing the % cell density infiltrating the IE region / the total infiltration for each tumor core, representing the specific cell subtypes indicated. Statistically significant p-values are noted atop each pair-wise comparison determined by one-way ANOVA. *, p<0.05; ****, p<0.001.

CD16a^high^ and CD16a^low^ NK cells infiltrated 95% and 90% of HGSC cores, respectively. In agreement with previous studies[28, 29], we found the largest frequency of tumor infiltrating immune cells to be macrophages (median = 149 cells/mm^2^) followed by T cells (median = 130.5 cells/mm^2^), while CD16a^high^ NK cells (median = 60.35 cell/mm^2^) and CD16a^low^ NK cells (median = 11.20 cell/mm^2^) infiltrated less frequently (Figure 1C). Although NK cells were represented at the lowest frequencies in both IS and IE regions, both CD16a^high^ and CD16a^low^ NK cells were overrepresented in IE regions, suggesting a preferential recruitment to engage with HGSC cells (p<0.05 compared with all other subsets; Figure 1D).

We next investigated how the frequency of immune cell infiltration associated with clinical parameters including patient age, *BRCA* status, tumor grade, tumor stage, and chemotherapy status (platinum-sensitive or resistant at 6 months post therapy) (Figure 2). There was no associateion between any immune cell infiltration in the IS or IE regions with *BRCA* or chemotherapy status, but significant associations were found with tumor stage and grade (Figure 2A,B). Patients with stage 4 HGSC exhibited higher CD16a^high^ frequencies in the IS region than patients with stage 3 HGSC (Figure 2C). Significantly more immune cells, including CD16a^high^ and CD16a^low^ NK cells, infiltrated grade 3 tumors compared with grade 2 tumors, in both the stromal and epithelial regions (Figure 2D). Similar significant increases in infiltration at higher grades were seen for IS T cells and macrophages, but no difference was observed in corresponding IE infiltrates (Figure 2D).

**Figure 2:**
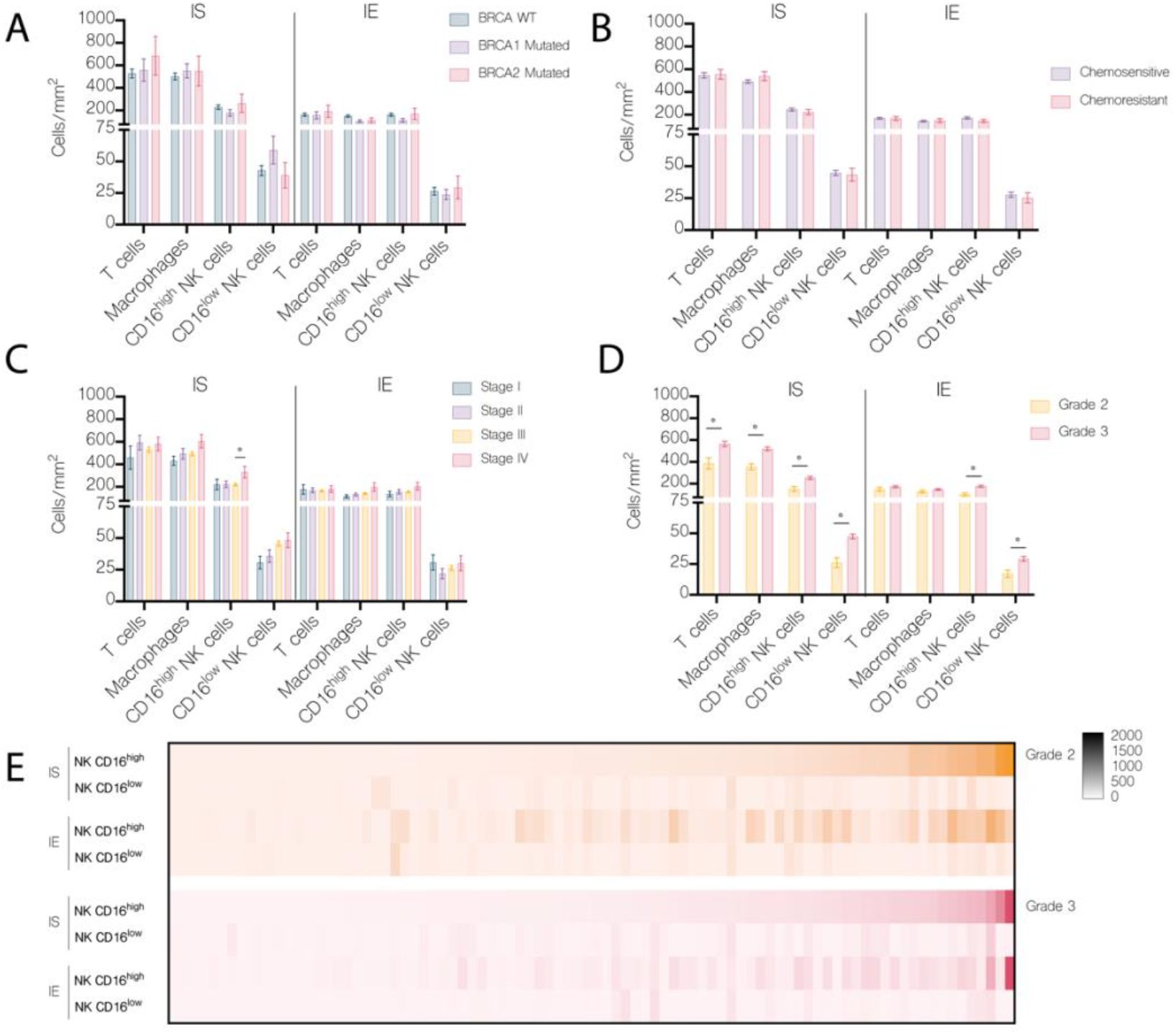
Immune cell infiltration into HGSC tumors stratified by BRCA status, grade, stage or chemosensitivity status. Cell density of each immune population in the intrastromal (IS) vs. intraepithelial (IE) tumor region stratified by A) BRCA status, B) chemotherapy status at 6 months C) tumor stage or D) tumor grade. E) Heat map visualizing infiltration by cell density of both NK cell populations, CD16a^high^ and CD16a^low^, in Grade 2 or Grade 3 HGSC tumors. Statistically significant p-values are noted atop each pair-wise comparison by cell type determined by either one-way ANOVA (3 or more comparisons BRCA status and tumor stage) or t-test (2 comparisons, tumor grade). *, p<0.05.

### IS CD16a^high^ NK cells predict improved 5-year progression free survival; IS CD16a^low^ NK cell predict poorer 5-year progression free survival

We conducted Kaplan-Meier analyses on the total, IE or IS immune cell infiltration using the median cell number to divide patients into groups of “high” and “low” infiltration. In line with previous studies in HGSC [28, 29], total T cells or total macrophages, without consideration for specific phenotype or inference of function (i.e. cytotoxic or regulatory; M1/M2), demonstrated no association with survival (Figure 3A). Interestingly, NK cells infiltrating into the IS region demonstrated survival associations dependent on their phenotype. Patients with ≥86 CD16a^high^ NK cells/mm^2^ in the IS region trended toward improved 5-year progression free survival (p=0.12); those with ≥19.5 CD16a^low^ NK cells/mm^2^ exhibited significantly poorer 5-year progression free survival (p=0.028) (Figure 3A). We reasoned that if the number of infiltrating NK cells predicted survival, then the top and bottom quartiles should be the most disparate with respect to their ability to predict survival outcomes. Indeed, comparing the quartiles in this way further distinguished the impact of NK cell infiltration: individuals representing top-quartile CD16a^high^ IS NK cell infiltration demonstrated significantly improved progression free survival compared to those with bottom-quartile CD16a^high^ infiltration (p=0.0471, Figure 3B).

**Figure 3:**
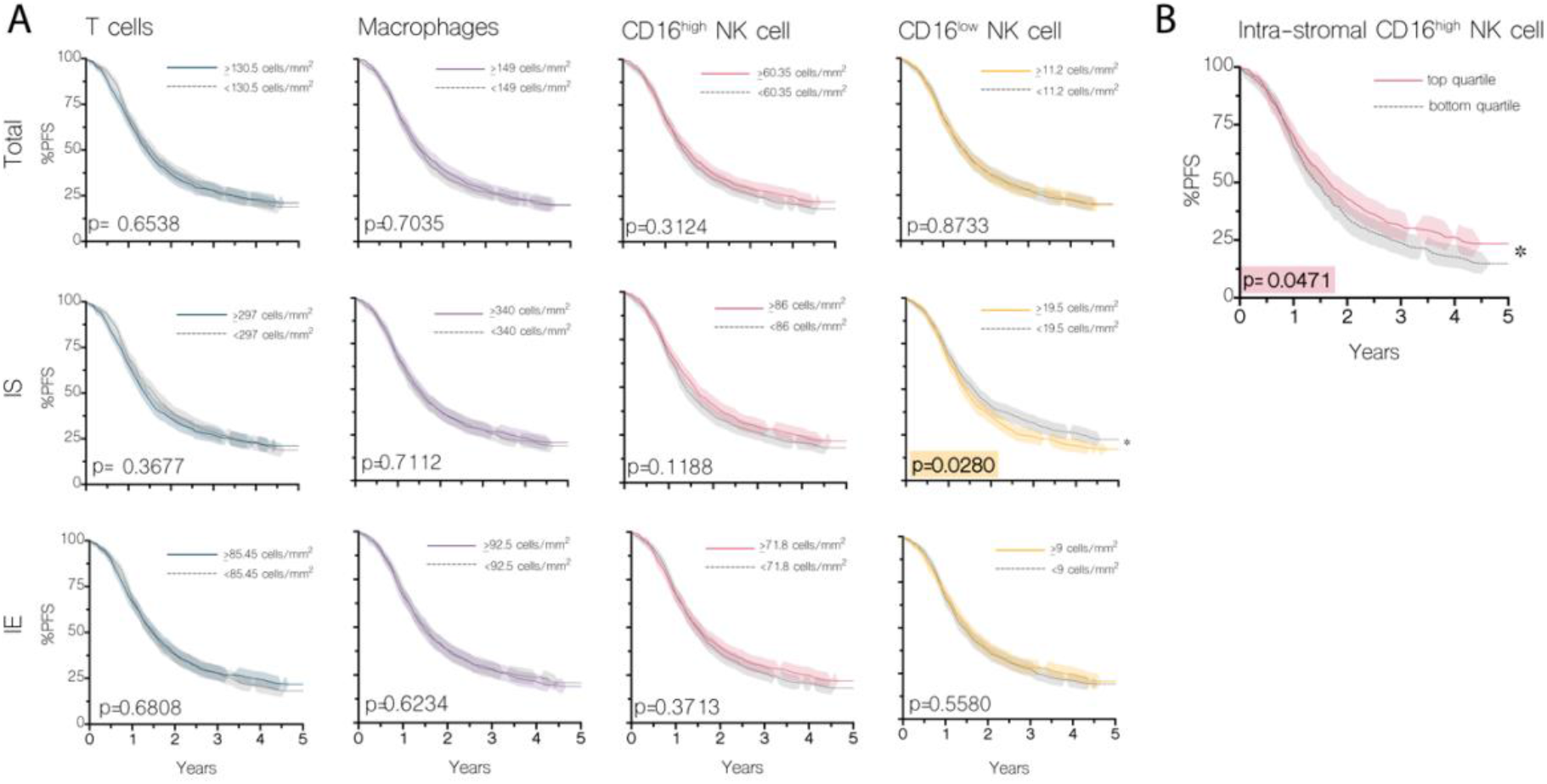
Kaplan-Meier survival curves for HGSC infiltration of total, IS and IE immune cells. A) The median density of immune cell infiltrates (cells/mm^2^) were used to dichotomize populations into either high or low immune infiltrate groups. Each immune cell population was assessed by infiltration to the tumor, or specifically to the IS or IE regions. B) To further investigate the benefit of CD16a^high^ NK cell infiltration, the population was broken into quartiles. Here the lowest and highest quartile were compared by Kaplan-Meier survival. Survival curves are surrounded by their 95% confidence intervals, with statistical significance tested by log-rank method.

### T cells, macrophages, and CD16a^high^ NK cells demonstrate shared patterns of co-infiltration, distinct from infiltrating CD16a^low^ NK cells

Co-infiltrating leukocytes can impact immune responses within HGSC tumors with influences on survival and treatment outcomes[30]. We next explored leukocyte co-infiltration into HGSC tumors generally, and within IS and IE tumor compartments. Most commonly, lymphocytes co-infiltrated the IE and IS with cells of the same subtype (T cells: r = 0.62; CD16a^high^ NK cells: r = 0.74; CD16a^low^NK cells: r = 0.48). The highest co-infiltration rates into the IS and IE regions were observed when comparing the values of infiltration of one lymphocyte type against itself: CD16a^high^ NK cells (r = 0.74); T cells (r = 0.62); and CD16a^low^ NK cells (r = 0.48) (Figure 4A). Macrophages, however, most commonly co-infiltrated with CD16a^high^ NK cells; most noteworthy was the correlation between IS macrophages and IS CD16a^high^ NK cells (r = 0.47) (Figure 4A). Interestingly, within each tumor compartment, co-infiltration between the two NK cell phenotypes was not frequently observed (IS; r = 0.31, IE; r = 0.22) (Figure 4A). In fact, co-infiltration between CD16a^high^ NK cells and either T cells or macrophages was more likely than co-infiltration of CD16a^high^ NK cells and CD16a^low^ NK cells in both IS and IE regions. Co-infiltration of CD16a^low^ NK cells and T cells and/or macrophages was rare in both regions. This indicates that tumors may exhibit patterns of infiltration that can be classified as “adaptive”, which consist of CD16a^high^ NK cells, T cells and macrophages or “innate” infiltrates which consists of CD16a^low^ NK cells, with or without T cells and macrophages (Figure 4B).

**Figure 4:**
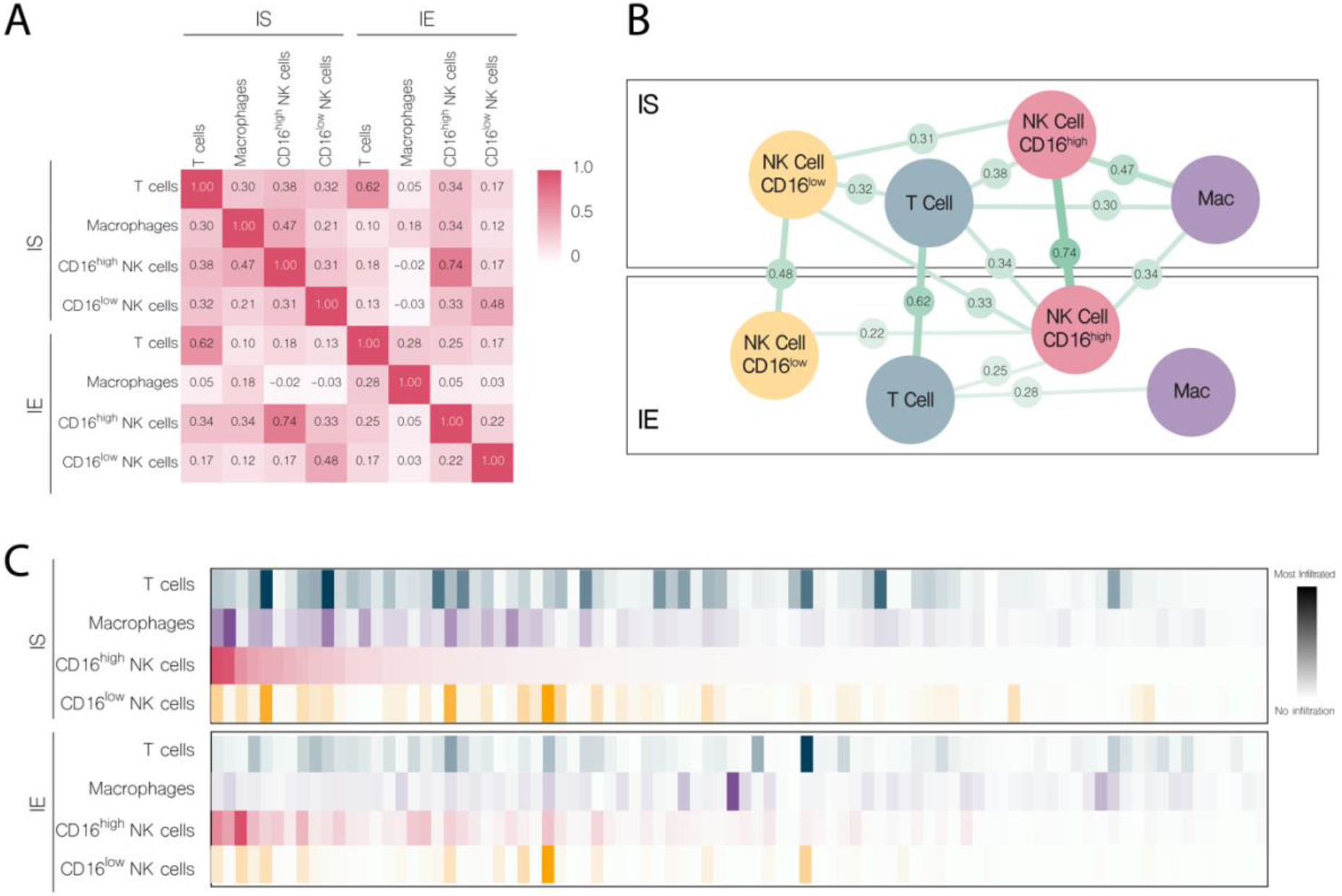
Co-infiltration between total, IS and IE immune cells demonstrates two independent patterns of immune cell infiltration: “adaptive” consisting of CD16a^high^ NK cells, T cells and macrophages, and “innate” consisting of CD16a^low^ NK cells. A) Pearson’s correlation matrix identifying co-infiltration of specific immune populations into each tumor region; IS and IE. B) Visualization demonstrating the relationship between infiltrating immune populations in in and between each tumor regions. The Pearson’s correlation values are represented as the lines between immune cell populations, with thickness of the lines representing the strength of those co-infiltration values. C) Heat map of 100 randomly selected, representative tumor cores with intrastromal and intraepithelial densities of each immune phenotype.

### Spatial analysis reveals that adaptive immune infiltrates predict longer progression free survival

We next asked whether the adaptive or innate infiltrate patterns predicted patient outcomes and used spatial analysis to define cellular “neighborhoods”. These neighborhoods were quantified based on the proximity of defined immune cells at a maximum of 22µm away from each other (Figure 5). Each neighbourhood was defined as either adaptive (one T cell, macrophage and CD16a^high^ NK cell), innate (two CD16a^low^ NK cells with or without other immune cells) or general (any other combination of at least three immune cells) (Figure 5A). These neighborhoods were then quantified based on the fraction of cells within the core that fell within them (the area of the neighborhood). We asked whether the neighborhood composition of cores mattered. While almost every core (98.2%) had some proportion of cells falling within the general neighborhood, 70% and 60% contained adaptive and innate neighborhoods, respectively (Figure 5B). When evaluating the prognostic potential of each, we observed that the frequency of general or innate cell neighborhoods did not associate significantly with progression free survival (Figure 5C). The presence of an adaptive neighborhood, however, was associated with a trend towards improved 5-year progression free survival (p = 0.069) (Figure 5C).

**Figure 5:**
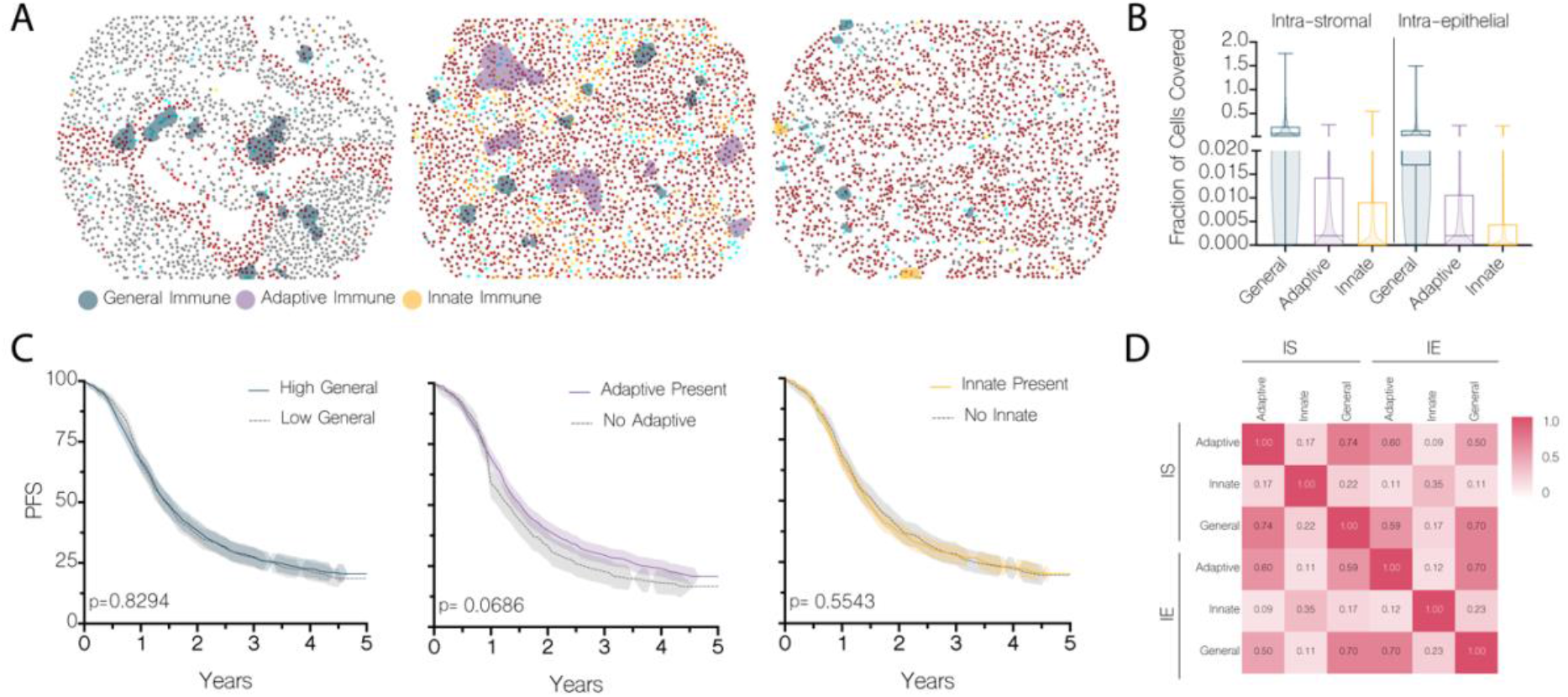
Adaptive immune neighborhoods of macrophages, T cells and CD16a^high^ NK cells predict longer progression free survival. A) Coordinates of each immune cell phenotype, identified by quantitative pathology, were imported to MatLab. Thereafter, cellular neighborhoods were identified by proximity at a maximum of 22µm distances. Neighborhoods were classified as adaptive immune (macrophages, T cells and CD16a^high^ NK cells), innate immune (a minimum of two CD16a^low^ NK cells), or general immune (any three immune cells). B) Superimposed bar graph and violin plot depict the frequency and distribution of fraction of cells covered by general, adaptive or innate neighborhoods. C) Immune cell neighborhoods were dichotomized above or below the median by either high vs low (general immune) or by the presence or absence of immune neighborhoods (adaptive or innate immunity). Differences in survival between groups was statistically evaluated by log-rank test. D) Pearson’s R correlation matrix of immune neighborhoods infiltrating within cores by tumor location

We next tested whether different types of immune neighbourhoods are present within the same cores. We found that the area of both adaptive and general neighborhoods was correlated in both the IS (r = 0.74) and IE (r = 0.70) regions of the same core (Figure 5D). In contrast, the area of innate neighborhoods was not correlated with that of general neighborhoods in the IS (r = 0.22) or IE (r = 0.23) regions, or with the area of adaptive neighborhoods in the IS (r = 0.17) or IE (r = 0.12). We conclude that immune cell infiltration is a general property of HGSC tumors, with skewing toward innate or adaptive phenotypes.

To determine whether the immunologic skewing might reflect tumor characteristics, we compared the immunologic phenotypes between patients based on tumor grade, stage, *BRCA* status and chemotherapy sensitivity. Both *BRCA1* and *BRCA2*-mutated tumors had significantly higher coverage by adaptive immune neighborhoods in the IE region (Figure 6A). While not statistically significant, we noted a trend toward higher coverage by innate neighborhoods in the IS region of chemotherapy resistant tumors (p = 0.074, Figure 6B). Similar to individual immune infiltration findings, we found a significant increase in the area covered by all immune neighborhoods in higher grade tumors (grade 3 vs. grade 2) (Figure 6D). Taken together, these findings suggest that the clinical or genetic characteristics of a tumor may predict its immune landscape.

**Figure 6:**
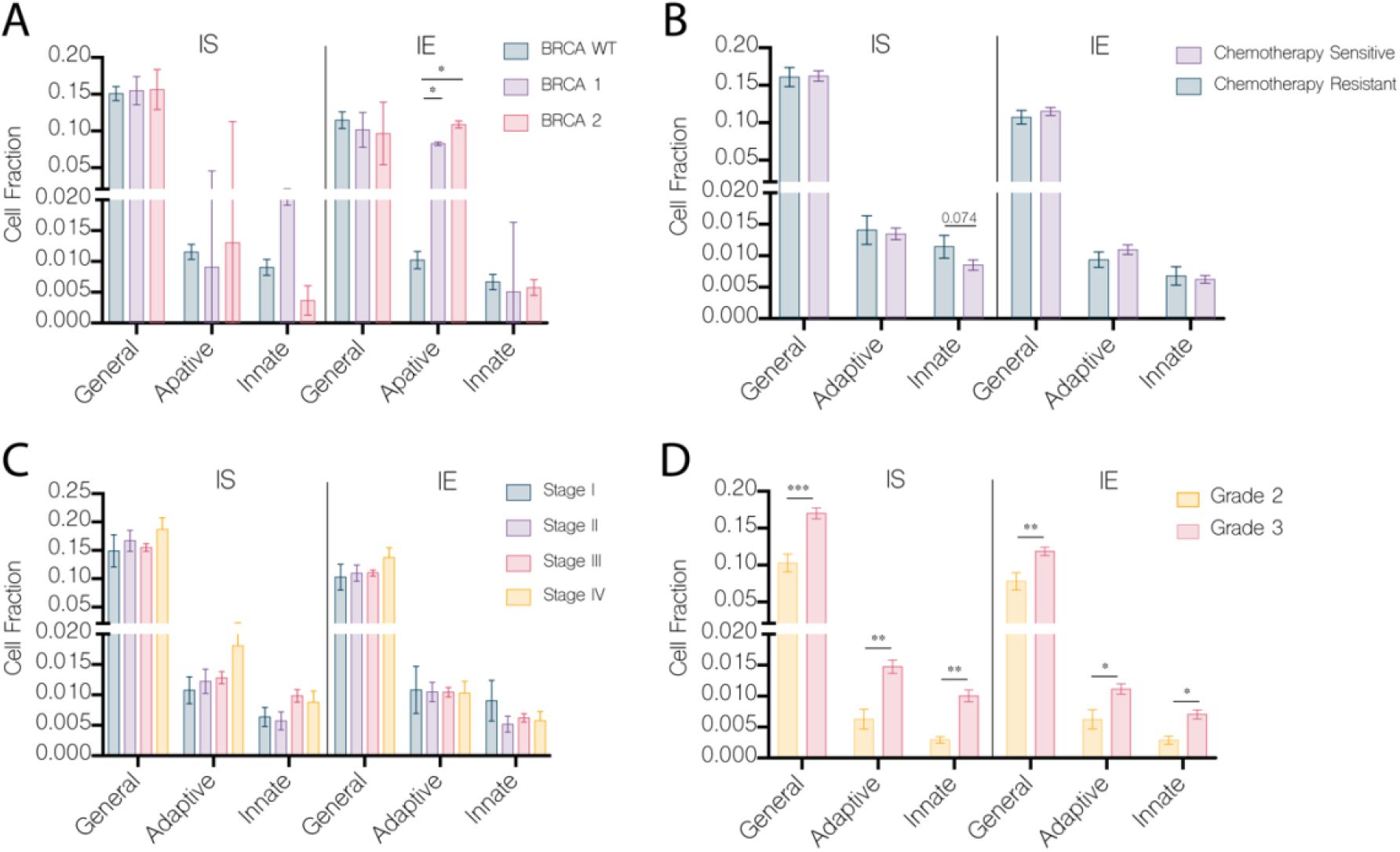
Adaptive immune neighborhoods are found at a higher frequency in BRCA1/2 mutated tumors, while innate neighborhoods are found in higher frequency in chemoresistant tumors. A) Cell density of each immune population in the IS vs. IE tumoral region stratified by BRCA status. B) chemotherapy status at 6 months C) tumor stage or D) tumor grade. Statistically significant p-values are noted atop each pair-wise comparison by cell type determined by either one-way ANOVA (3 or more comparisons BRCA status and tumor stage) or t-test (2 comparisons, tumor grade). *, p<0.05.

### CD16^high^ and CD16^low^ NK cells are dichotomized by expression of CD39/CD73

Our TMA analysis demonstrates that CD16^high^ and CD16^low^ NK cells have opposite associations with prognosis for patients with HGSC. To better understand the features of these two populations, we performed flow cytometry on PBMCs from six healthy donors challenged with ovarian cancer cell lines (OVCAR4 and OVCAR5) or the HLA-negative NK cell target, K562. We compared both populations for markers of activation and inhibition, as well as expression of CD39/73, exonucleosidases associated with degradation of ATP to adenosine, establishment of an immunosuppressive microenvironment in HGSC and regulatory NK cell function[31-33]. Consistent with the missing self-responsiveness expected against an HLA-negative target cell, the greatest responsiveness against K562 cells was mediated by CD16^bright^ NK cells (which are enriched for the educated NK cell subset (Figure 7A). However, OVCAR4 and OVCAR5 cells, which persist in HLA expression, were targeted by both CD16a^high^ and CD16a^low^ subsets of NK cells.

**Figure 7:**
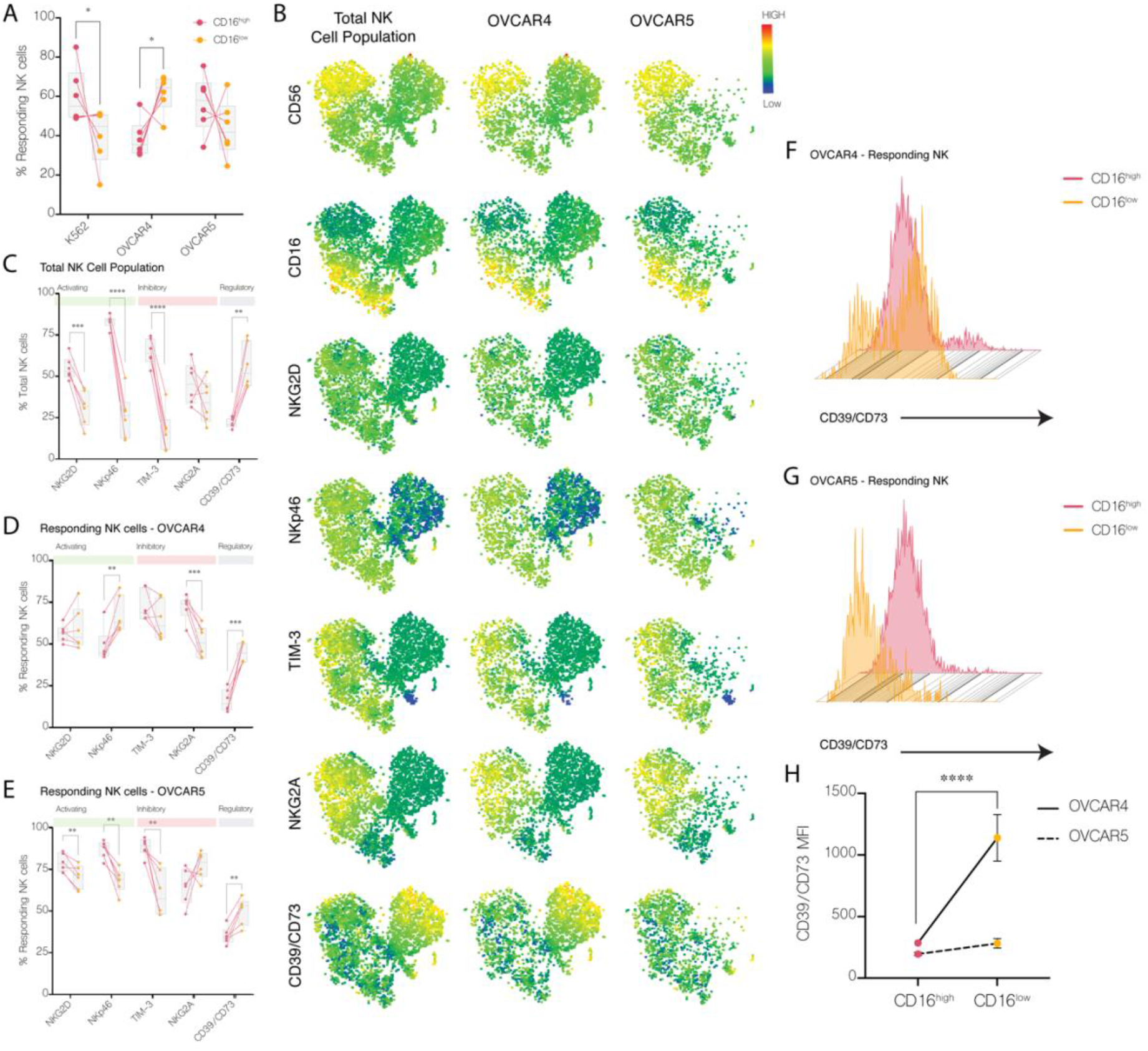
CD16^high^ NK cells demonstrate a phenotype consistent with immunoregulation. Flow cytometry analysis was conducted on NK cells co-cultured with K562, OVCAR4 and OVCAR5 cell lines. A) The percent of responding NK cells in each co-culture comparing CD16^high^ to CD16^low^ NK cells. Lines join populations from each independent donor. B) t-SNE representation of CD56, CD16 and CD39/CD73 from the combined NK cell populations responding to target cell stimulation, or NK cell populations responding against OVCAR4 and OVCAR5 cells. C) Percentage of total or responding NK cell populations positive for phenotypic markers NKG2D, NKp46, TIM-3, NKG2A and CD39/73 of NK cells from each donor (represented by an individual dot with CD16^high^and CD16^low^ NK cells from the same donor joined by a line) and those responding in co-cultures with D) OVCAR4 or E) OVCAR5 cell lines. Histograms illustrating CD39/CD73 expression of responding NK cell population against F) OVCAR4 and G) OVCAR5 cell lines and G) corresponding MFI quantification. Statistically significant p-values are noted atop each pair-wise comparison by cell type determined by paired t-tests. ****, p<0.001 (OVCAR4).

Compared with responding CD16a^dim^ NK cells, responding CD16a^bright^ NK cells expressed higher NKp46 and NKG2D activating receptors, and the TIM-3 inhibitory receptor. Noteworthy, the responding CD16a^dim^ population was consistently enriched for cells expressing CD39/CD73 compared with the responding CD16^bright^ population (Figure 7B – G), suggesting that the C16^dim^ may have immune regulatory potential in the ATP/adenosine-rich tumor microenvironment.

## DISCUSSION

To our knowledge, this is the most in-depth and specific exploration of NK cell infiltration into HGSC. Using a TMA that represented 1145 patients with HGSC, we have refined and expanded the predictive prognostic value of NK cell infiltration into HGSC tumors. Using multiplex IF, and pathologist-validated machine learning, we spatially resolved all cells with an NK cell phenotype, and differentiated subsets based on CD16a staining intensity. We report that both the subtype of NK cell and its localization within the TME contribute to prognostic associations for patients with HGSC. The biggest impacts of cellular infiltration on progression free survival were observed among NK cell subsets infiltrating the IS region: infiltration of CD16a^low^ NK cells is associated with poor progression free survival; infiltration of CD16a^high^ NK cells is associated with improved progression free survival. Using a correlational approach, we find that CD16a^high^ NK cells co-infiltrate with T cells and macrophages; hence the presence of CD16^high^ NK cells might predict for infiltration of a collection of immune cells that contribute to control of HGSC.

The function of NK cells, defined by co-expression of germline-encoded receptors, remains to be defined in HGSC, but the dichotomy of prognostic impacts that associate with CD16a phenotypes points to differential function among NK cell subsets[34]. Typically, CD16a^high^ NK cells are the most frequent in the blood, have high cytotoxic function, KIR expression, education for missing self-reactivity and produce pro-inflammatory cytokines. CD16a^low^ NK cells, by contrast, can take on an array of functions including cytokine production and immune regulation[16, 35]. Among this CD16^low^ subset, we identified a high frequency of cells expressing CD39/73. These are ectonucleotidases associated with degradation of ATP to AMP, then ADO, and have been associated with immunoregulatory functions among NK and other immune cells[32]. High expression of CD73 and CD39 associates with the immunoregulatory subtype of HGSC[32], and its associated poor prognosis[33]. CD39/73^+^ NK cells were responsive in our *in vitro* analysis, but this was conducted in the absence of the immunosuppressive and adenosine-rich microenvironment of HGSC. For CD16a^dim^CD39/73^+^ NK cells, the tumor microenvironment would be expected to provide signals for adenosine-mediated inhibition and potentially interrupt cytotoxic NK cell function. The function of CD16^dim^CD39/73^+^ NK cells may be interrupted, or become immunoregulatory, in the tumor microenvironment, which may account for its association with relatively poorer outcomes for patients.

The impacts of tumour-infiltrating NK cells in HGSC are controversial, with studies alternately ascribing beneficial or detrimental roles. This may reflect the way that NK cells were appraised in these studies. In a recent meta-analysis, we demonstrated that studies across all solid tumors were more likely to identify a positive prognostic association of NK cells if they were stained using antibodies against CD56 or CD57 rather than NKp46[36]. CD56 staining is unreliable in HGSC because the tumor cells themselves frequently (∼60%) express CD56[20]. CD57 expression is likewise not ideal: though restricted to NK cells, it is selectively expressed on the CD56^dim^CD16a^high^ population. Indeed, studies of HGSC that defined NK cells using CD57 staining have defined a prognostic benefit of NK cell staining [24, 25]; likely, this reflected measurement the CD16^high^ NK cell population. Staining with NKp46 would capture a broader population of NK cells, but also innate-like lymphocytes (ILCs), which are known to have immunosuppressive activity that interferes with immune-mediated tumor cell killing[37, 38]. To circumvent these challenges while specifically and completely marking NK cells, we stained for CD94 (a component of the binding complexes formed by the NK receptor group 2 (NKG2) receptors found on all NK cells, validating this approach using PBMC, tonsil samples and HGSC cores on which CD56 epithelial staining did not occur (data not shown).

An additional feature that allowed us to refine NK cell associations was our division of the IS and IE regions by including pancytokeratin staining in our multiplex panel[24, 25]. With this approach, we identified that NK cells are more likely to be recruited to the IE than T cells or macrophages, and that the prognostic benefit of CD16a^high^ and detriment of CD16a^low^ NK cells was associated with their frequency in the IS regions specifically. Within the IE, NK cells would be exposed to inhibitory ligands and checkpoints, including NKG2A[20]; nevertheless, the localization of NK cells to the epithelial region supports strategies to maximize NK cell reactivity or recruit these cells from the neighboring IS regions. For patients lacking CD16a^high^ NK cells, strategies to inflame the tumor and induce an infiltration of leukocytes with anti-tumor function may be required to facilitate tumor control[39, 40]. Further, a key consideration is how tumour progression and treatment may influence the state of immunity within the TME. We have previously reviewed the many additional alterations found in NK cells isolated from ovarian cancer patient tumors and ascites elsewhere[41]. For example, NK cells isolated from the ascites of ovarian carcinoma patients had significantly decreased CD16a expression compared to that found in peripheral blood[42].

Conclusive functional studies are beyond the scope of this report, but our observations suggest that there is a setpoint for immunity that differs between patients with ovarian HGSC. Both CD16^high^ and CD16^low^ NK cells can be recruited to sites of infection or cancer by chemokines and influenced by local cytokines in the TME, which may differ between patients[39, 43]. We found that although the types of infiltrating NK cells were not mutually exclusive, the majority of tumor cores that we studied were preferentially infiltrated by one subtype. “Adaptive” immune cell neighborhoods, defined by the presence of CD16a^high^ NK cells, T cells and macrophages, were most often found in cores with other adaptive and “general” immune cell neighborhoods, but not with “innate” neighborhoods, which consisted of CD16a^low^ NK cells. Innate neighborhoods, by contrast, were small or absent in cores with adaptive neighborhoods representing larger areas. We identified associations with clinical features: the presence of adaptive clusters was associated with *BRCA* mutations and higher-grade tumors, while the presence of innate clusters was associated with chemotherapy resistance. Our samples were all taken from treatment-naïve patients that went on to receive therapy. Perhaps the availability of CD16^high^ NK cells, T cells and macrophages in patients with adaptive clusters fosters productive antitumor immunity by collaborating on key features such as direct immunogenic cell death facilitated by NK cell cytotoxicity, followed by antigen presentation and cytokine polarization[44]. Those with innate clusters, defined by CD16a^low^ NK cells, may instead be associated with a more suppressive TME (i.e. TGF-β, IL-10 checkpoints, CD39/73 expression on NK and other cells[33]), and/or the lack of anti-tumor effector cells.

Ovarian HGSC tumors carry a high neoantigen burden, which has made them candidates for immunotherapy. However, checkpoint inhibition strategies have not impacted HGSC substantially [4-6], suggesting a failure of lymphocytes to completely recognize and control tumors. Indeed, sequencing of the TCRs of tumor-infiltrating T cells demonstrated that the majority do not have specificity for tumor antigens[45]. Moreover, HGSC progression, genomic instability and checkpoint therapy can promote outgrowth of mutants with loss-of-heterozygosity or disruption of components of the HLA processing and presentation pathway, which enables escape from T cell mediated (i.e. HLA-restricted) recognition[46, 47]. With these features in mind, the roles of infiltrating NK cells may be to initiate tumor killing via antigen-independent mechanisms, promote phagocytosis and presentation of neoantigens to prime T cells, and missing self-recognition and killing of HLA-negative tumors. These features are consistent with NK cells expressing high levels of CD16a[16]. For this study, we chose to focus on NK cells and their localization within tumor regions. Limitations in the staining panel size meant that we were unable to study the function of macrophages (i.e. discern as suppressive M1 or pro-inflammatory M2) or T cells (as CD4, CD8, regulatory), but previous studies have revealed that infiltration of cytotoxic CD8^+^ T cells or M1-polarized macrophages are independently associated with improved overall survival in patients with HGSC[48, 49]. Acting together with C16^high^ NK cells, these anti-tumor leukocytes might best respond to the diverse array of phenotypes of HGSC cells that occur. Our current findings endorse strategies to recruit or active CD16a^high^ NK cells in immunotherapies for HGSC. Future studies to clarify the key features, functions and interactions between NK cells and other leukocytes may inform immunotherapies to best recruit CD16^high^ NK cells to maximize HGSC control.

## Data Availability

Aggregate clinical data is listed in supplementary table 1.
Quantitative pathological cell density and spatial pathology quantification by core is available upon request.

## DECLARATIONS

### Ethics

All methods for specimens and clinical information collection and subsequent analyses were approved by Dalhousie University’s Research Ethics Board (#2020-5060). Primary PBMC used to validate antibodies were approved by the Dalhousie University REB (#2016-3842) and the Canadian Blood Services REB (#2016-016).

### Consent for Publication

All authors consent to publication of this work.

### Availability of data and material

Aggregate clinical data is listed in supplementary table 1.

Quantitative pathological cell density and spatial pathology quantification by core is available upon request.

### Author’s Contributions

The studies were conceptualized by SN, BHN and JEB; experiments were designed and validated by SN, SNL, SG, LM, LC, BHN, AMMM and JEB. Pathology review and validation of machine learning and analysis was conducted by SN and TA. Computational spatial analysis was designed by SN, DA and JEB, and conducted by DA. Flow cytometry and functional analysis was conducted by SNL. The manuscript was written by SN, SG and JEB, reviewed, edited, and approved by all authors.

## Acknowledgements

The authors gratefully acknowledge experimental assistance from Ms. Morgan Pugh-Toole and Katy Milne, and PBMCs provided in collaboration with Canadian Blood Services. This study uses resources provided by the Canadian Ovarian Cancer Research Consortium’s -COEUR biobank funded by the Terry Fox Research Institute and Ovarian Cancer Canada and managed and supervised by the Centre de recherche du Centre hospitalier de l’Université de Montréal (CRCHUM). The Consortium acknowledges contributions to its COEUR biobank from Institutions across Canada (for a full list see www.tfri.ca/COEUR/members).

## List of Abbreviations

COEUR: The Canadian Ovarian Experimental Unified Resource
FFPE: formalin-fixed paraffin-embedded
HGSC: high grade serous carcinoma
IE: Intraepithelial
IS: Intrastromal
HLA: human leukocyte antigen
NK: natural killer
PBMC: peripheral blood mononuclear cells
TME: tumour microenvironment

