## Supplemental Material for "CD16a^high^ NK cell infiltration and spatial relationships with T cells and macrophages can predict improved progression-free survival in high grade ovarian cancer"

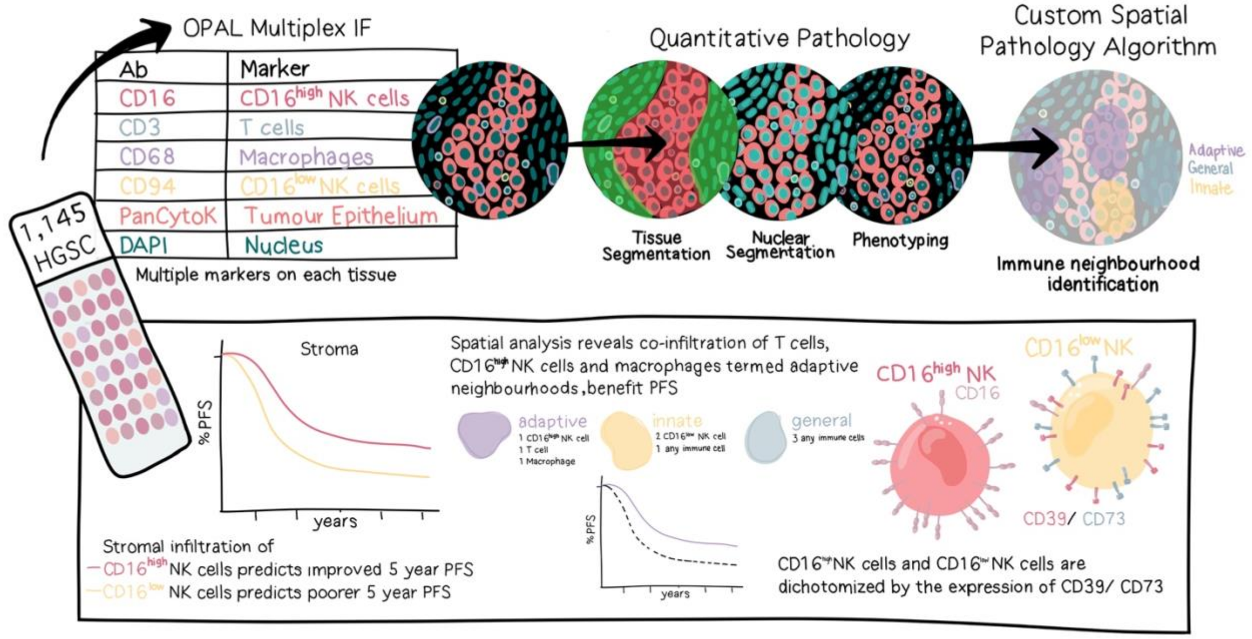


**Supplemental Figure 1: Graphical abstract.**

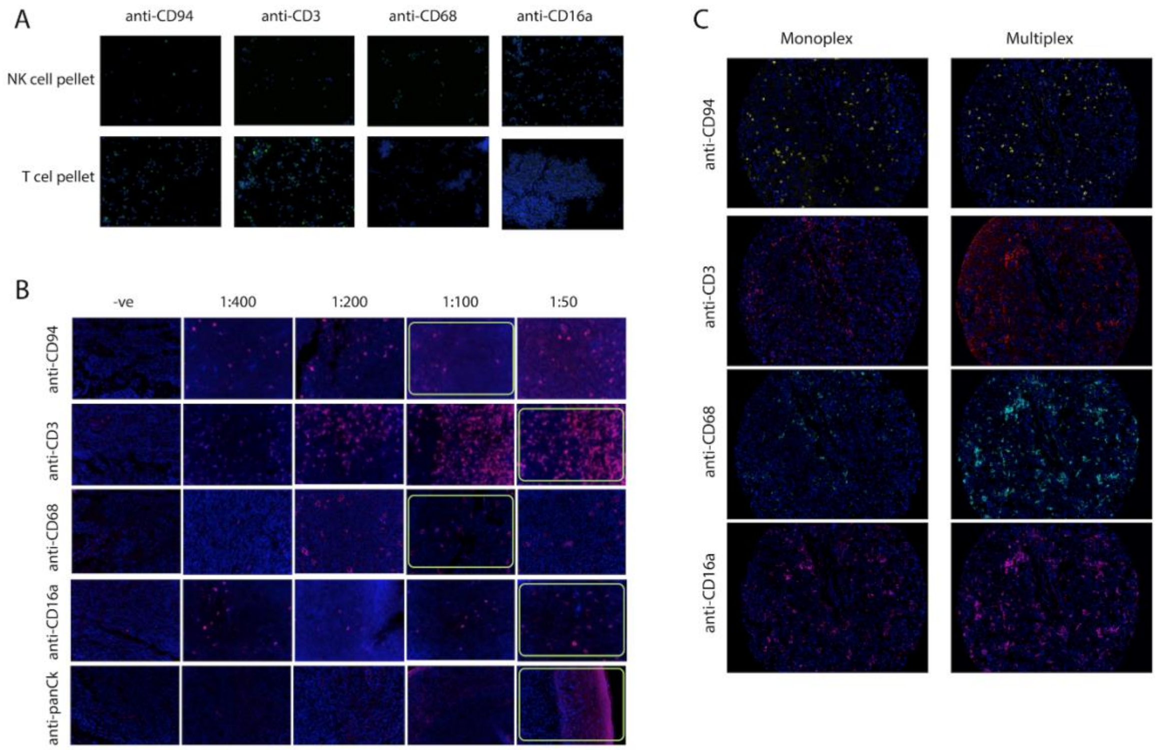


**Supplemental Figure 2**: **Antibody and staining validation.** A) NK and T cell pellets for positive/ negative validation. B) Antibody dilutions on tonsil tissue. Green boxes indicate the concentration used for multiplex. C) Monoplex/Multiplex staining on HGSC optimization tissue. Image captured on the Mantra (Akoya Biosciences) at a magnification of 20x.

| Tissue | PanCk Pathology Views | InForm Segmentation | Pathologist Segmentation | DAPI Pathology Views | InForm Cell Segmentation | InForm Count | Pathologist Count | InForms accurarcy in % |
|---|---|---|---|---|---|---|---|---|
| 1 (C56) | 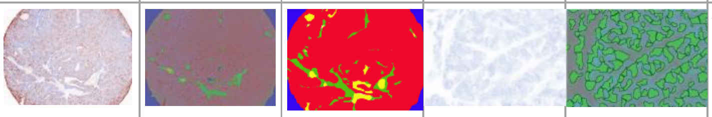 | | | | | 119 | 175 | 68% |
| 2 (F76) | 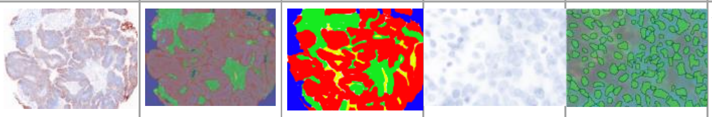 | | | | | 130 | 147 | 88% |
| 3 (E171) | 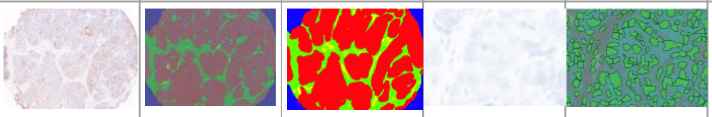 | | | | | 143 | 156 | 92% |
| 4 (C12) | 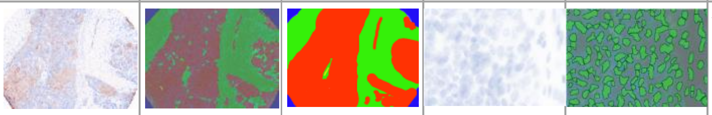 | | | | | 135 | 142 | 95% |
| 5 (C221) | 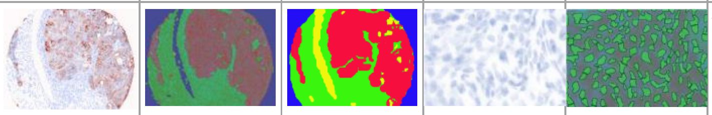 | | | | | 143 | 190 | 75% |
| 6 (C196) | 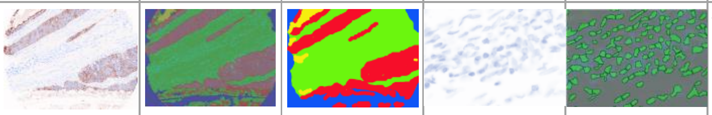 | | | | | 105 | 113 | 93% |
| 7 (C104) | 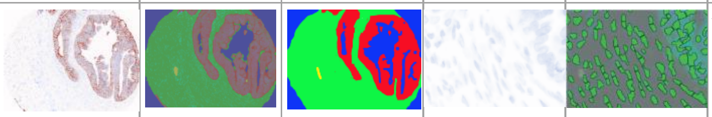 | | | | | 116 | 105 | 110% |
| 8 (C230) | 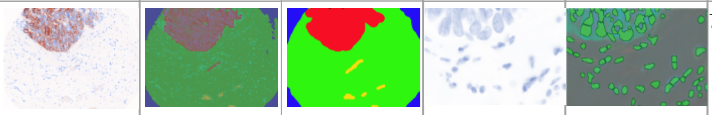 | | | | | 71 | 60 | 118% |
| 9 (C162) | 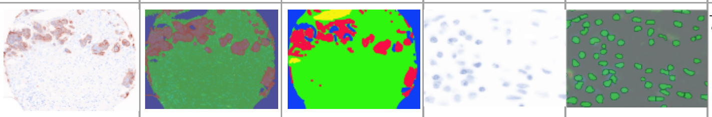 | | | | | 70 | 67 | 104% |
| 10 (E272) | 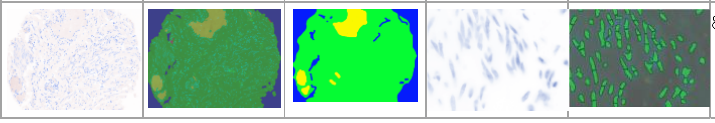 | | | | | 83 | 73 | 113% |

**Supplemental Figure 3: Representative digital and manual pathology segmentation and scoring.** Tissue segmentation was assigned to each core after training of the machine learning algorithm in InForm digital pathology software. Pathologist TA segmented the same cores, blinded to the InForm outcomes. Pathology views, InForm segmentation and the pathologist's segmentation are shown. Cell segmentation was trained to recognize individual cell nuclei and InForm or pathologist TA independently counted individual cells. The range and variation between counts is shown.

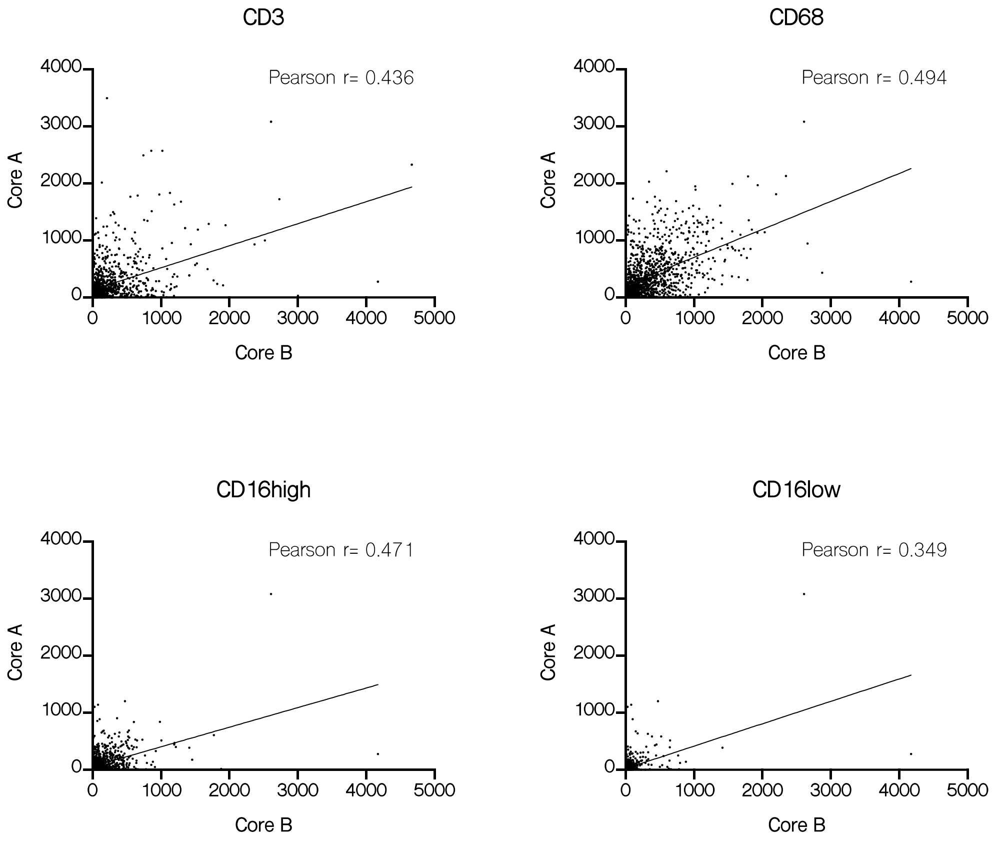


**Supplemental Figure 4: Pearson correlation between cell density quantified in duplicate cores (Core A vs. Core B) isolated from the same patient tumourss.**

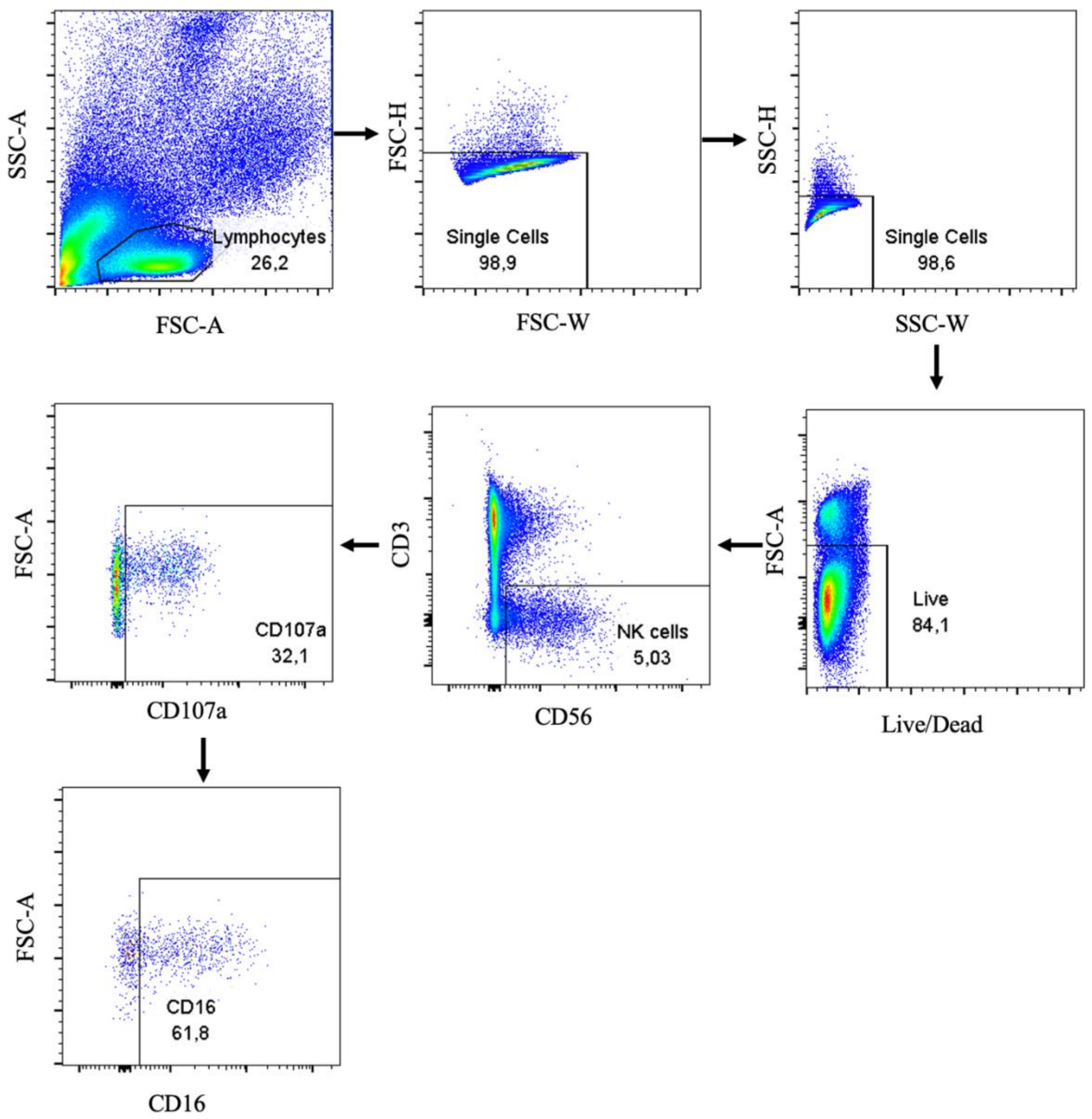


**Supplementary Figure 5. Gating strategy.**

**Supplementary Table 1: Aggregate clinical data.**

| Characteristics | Category | N (%) |
|---|---|---|
| Patients | Number of patients represented in TMA | 1,145 (100%) |
| Age | Median (range) | 62 (26-91) |
| Diagnosis Year | 1992-1995 | 14 (1.2%) |
| | 1996-2000 | 104 (9.1%) |
| | 2001-2005 | 302 (26.4%) |
| | 2006-2010 | 482 (42.1%) |
| | 2011-2012 | 169 (14.8%) |
| | 2013-2014 | 49 (4.3%) |
| Tumour Grade | Grade 2 | 132 (11.5%) |
| | Grade 3 | 850 (74.2%) |
| FIGO Tumour Stage | I | 76 (6.6%) |
| | II | 120 (10.5%) |
| | III | 789 (68.9%) |
| | IV | 107 (9.3%) |
| BRCA Status | BRCA WT | 330 (28.8%) |
| | BRCA1 mutated | 52 (4.5%) |
| | BRCA2 mutated | 19 (1.7%) |
| | BRCA1/2 mutated | 2 (0.2%) |
| Debulking Status | No residual disease | 277 (24.2%) |
| | Residual disease | 584 (51%) |

**Supplementary Table 2: OPAL TSA-based multiplex immunofluorescence panel.**

| **Marker** | **Round** | **Species** | **Clone #** | **Blocking** | **AR** | **Polymer** | **Ab Dilution** | **Fluorophore** | **OPAL Dilution** |
|---|---|---|---|---|---|---|---|---|---|
| CD3 | **1** | Mouse | LN10 | OPAL | 9 | Ms + Rb HRP | 1/50 | 650 | 1/100 |
| CD16a | **2** | Rabbit | SP175 | OPAL | 9 | Ms + Rb HRP | 1/50 | 520 | 1/100 |
| CD94 | **3** | Rabbit | ERP21003 | OPAL | 9 | Ms + Rb HRP | 1/100 | 570 | 1/100 |
| CD68 | **4** | Mouse | PGM1 | OPAL | 9 | Ms + Rb HRP | 1/100 | 620 | 1/100 |
| panCk | **5** | Mouse | AE1/AE3 | OPAL | 9 | Ms + Rb HRP | 1/50 | 480 | 1/50 |
| DAPI | **6** | | | | | | | | |

**Supplementary Table 3: Flow Cytometry antibody panel.**

| Antibody Target | Clone | Source |
|---|---|---|
| **CD3** | UCHT-1 | BD Biosciences, San Jose, CA |
| **CD56** | NCAM16.2 | BD Biosciences, San Jose, CA |
| **CD16** | 3G8 | BD Biosciences, San Jose, CA |
| **CD39** | Tü66 | BD Biosciences, San Jose, CA |
| **CD73** | AD2 | BD Biosciences, San Jose, CA |
| **TIM-3** | 7D3 | Biolegend, San Diego, CA |
| **NKG2A** | REA110 | Miltenyi Biotec, San Diego, CA |
| **NKG2D** | 1D11 | BD Biosciences, San Jose, CA |
| **NKp46** | 9-E2 | BD Biosciences, San Jose, CA |
| **CD107a** | H4A3 | BD Biosciences, San Jose, CA |
